# A Double-Blind, Sham-Controlled Multi-Site Pilot Randomized Trial of Amygdala tFUS in Treatment-Resistant Generalized Anxiety Disorder

**DOI:** 10.64898/2026.11.24.26368811

**Authors:** Norman M. Spivak, Andrew A.E.D. Bishay, Amber Hopkins, Andrew J. Swenson, Jonathan Haroon, Ashley Hayden, Elisabeth Collins, Samantha F. Schafer, Edythe D. London, Martin L. Lee, Alexander Bystritsky, Mark E. Schafer, Tina Chou, Kevin Caulfield, Darin Dougherty, Mark S. George, Susan Y. Bookheimer, Martin M. Monti, Margaret Distler

**Author notes:** Deceased (Edythe D. London). Served as Site PI. **Disclosures:** NSp, ABi report having received consulting fees from BrainSonix. ABy is the founder and CEO of BrainSonix. MSc & SSc are inventors on neuromodulation patents. All other authors report no financial relationships with commercial entities.

## Abstract

**Objective:** Transcranial focused ultrasound (tFUS) enables noninvasive, spatially precise modulation of deep brain structures, including the amygdala, a key node in anxiety-related circuitry. We evaluated whether repeated right-amygdala tFUS improves symptoms in patients with treatment-resistant generalized anxiety disorder (GAD).

**Methods:** In this multisite, double-blind, sham-controlled pilot trial, 31 adults with treatment-resistant GAD were randomized 1:1 to active or sham right-amygdala tFUS. Across three academic centers, participants received four sessions over one month and were followed for a month afterward. The primary endpoint was baseline-to-final-visit change in Hamilton Anxiety Rating Scale (HAM-A) score. Post hoc, subject-specific acoustic modeling assessed target engagement among active-treatment participants at UCLA.

**Results:** tFUS delivered both in the scanner (visits 1,4) and out of the scanner (visits 2,3) was well tolerated with no worsening of anxiety. HAM-A scores decreased by 10.33 points (SD 9.51) with active tFUS and 8.81 points (SD 9.89) with sham, with no significant between-group difference (mean difference, −1.52; 95% CI, −8.65 to 5.61; *p*=.333; Cohen’s *d*=−0.16). HAM-A scores declined significantly over time across participants (*p*<.001), but trajectories did not differ between groups (*p*=.934). Acoustic modeling in four active participants demonstrated overlap of the simulated field with the right amygdala, although target coverage was limited and variable.

**Conclusions:** Repeated right-amygdala tFUS was feasible for treatment-resistant GAD, but this small study found no significant clinical benefits over sham. Improvements in both groups, no differential physiological effects, and limited target coverage warrant adequately powered trials testing optimized dosing, precise targeting, and state-dependent or symptom-provocation paradigms.

## Introduction

Generalized anxiety disorder (GAD) is a prevalent (3–5%) and disabling condition characterized by excessive, persistent worry and substantial functional impairment^1^. Despite available treatments, as many as 40% of treatment-seeking patients remain symptomatic^2^, with response rates of approximately 50% for pharmacotherapy^3^ and 55% for cognitive behavioral therapy across anxiety disorders^4^. These limitations underscore the need for novel interventions for treatment-resistant GAD.

The amygdala plays a central role in threat processing and emotion regulation, with altered amygdala activity and connectivity implicated in GAD^5,6^. Neuroimaging studies have also identified abnormal right-amygdala connectivity in patients with anxiety^7^, providing a rationale for targeted neuromodulation.

Unlike conventional noninvasive brain stimulation techniques, which have limited access to deep brain structures, low-intensity transcranial focused ultrasound (tFUS) enables spatially focused neuromodulation of deep targets, including the amygdala^8 9^. In a previous open-label study of patients with treatment-resistant GAD, right-amygdala tFUS was associated with a >30% reduction in Hamilton Anxiety Rating Scale (HAM-A) scores in 60% of participants, with 32% achieving remission^10^. However, as anxiety has a high placebo rate (up to 50% in some pharmacologic studies ^11^), more stringent and blinded trials are needed. In the present study, we hypothesized that targeted tFUS to the right amygdala would reduce anxiety symptoms relative to sham stimulation. Here, we report the findings of a double-blind, sham-controlled randomized clinical trial of tFUS targeting the right amygdala in subjects with treatment-resistant GAD.

## Methods

The study was registered on ClinicalTrials.gov (NCT04557891) before the first patient was enrolled. Patients with treatment-resistant GAD were enrolled across 3 sites (University of California, Los Angeles (UCLA), Massachusetts General Hospital (MGH), and Medical University of South Carolina (MUSC)). UCLA served as the data coordinating center, and the UCLA Medical IRB reviewed the study (IRB: 20-1750); sites also maintained local IRB approvals. All participants provided written, signed informed consent after the study was explained and before any study activities began.

### Participants

Thirty-one patients (67.74% female) were enrolled in the study across the three sites. Participant age and sex were obtained by self-report. Race and ethnicity were not systematically collected as part of the study protocol. The lack of race and ethnicity data limits our ability to characterize the sample’s demographic diversity and assess the generalizability of the findings across racial and ethnic groups.

Patients were required to have a diagnosis of treatment-resistant GAD with no other comorbid diagnoses. A diagnosis of depression was permitted insofar as the depression was secondary to GAD. Treatment resistance was defined *a priori* as having failed two selective serotonin reuptake inhibitors (SSRIs) and one non-serotonergic augmentation (e.g., a noradrenergic-dopaminergic drug [bupropion] or eight sessions of CBT, etc.). This definition aligns with that of other device-based trials in psychiatric disorders^12^. Patients were required to be stable on medications for at least 1 month before study initiation. Participants were between the ages of 18 and 65 and were medically stable, with normal or corrected-to-normal vision and hearing. After eligibility was confirmed using the Mini International Neuropsychiatric Interview ^13^, patients were randomized in a 1:1 ratio to receive either active tFUS or sham stimulation. All participants completed a total of seven study visits, including MRI-based assessments at baseline (Visit 1) and following the final treatment session (Visit 4).

### Study Design

This was a double-blind, randomized, sham-controlled feasibility trial conducted across three academic medical centers. Participants underwent seven study visits over approximately two months. The intervention consisted of four sessions of tFUS or sham stimulation targeting the right amygdala, administered on visits 1 through 4. No tFUS was administered during visits 5 through 7. These were follow-up visits in which outcome measures were re-assessed, performed 1 day, 1 week, and 1 month, after visit 4, respectively. In visits 1 and 4, tFUS was administered inside an MRI machine, and included acquisition of resting-state functional neuroimaging before, during, and after sonication. In visits 2 and 3, tFUS was administered outside the MRI machine, and targeting was accomplished with frameless stereotactic neuronavigation (BrainSight, Rogue Research, Montreal, QC). Each treatment visit involved two administrations of tFUS, wherein each administration was 5 minutes of stimulation delivered over 10 minutes in a 30 second ON, 30 second OFF paradigm. A total of 10 minutes of stimulation was delivered for each visit. Randomization was stratified by site. All study personnel and participants were blinded to group allocation, designated as either group A or B. Only the study sponsor (BrainSonix Corp., Sherman Oaks, CA) held the key to A/B assignment, which was varied by site to enhance blinding integrity. Sham stimulation was delivered using BrainSonix’s patented (US Patent 12,201,856) sham pads, which are visually indistinguishable; one attenuates >99% of ultrasound energy, while the other permits near-complete transmission^14,15^.

### Imaging Procedures

The imaging protocol included high-resolution structural imaging consisting of a T1-weighted MPRAGE, which was used for neuronavigation targeting during visits 2 and 3 and in-scanner guidance during visits 1 and 4.

### Outcome Measures

The primary efficacy endpoint was the change in HAM-A score from baseline (Visit 1) to the final treatment visit (Visit 4), analyzed using between-group comparisons of change scores. This analysis was conducted using available-case data, including participants with complete data at both time points.

As a secondary analysis, longitudinal changes in HAM-A across all study visits were assessed using repeated measures ANOVA, incorporating all available observations. All participants with available post-baseline data were included.

### Stimulation Parameters

tFUS was delivered using a BrainSonix BX Pulsar 1002, using a 65mm focal depth transducer targeting the right amygdala^16^. Sonication was delivered at an I_SPPA.3_ of 14.4 W/cm^2^, a pulse repetition frequency of 10 Hz, a 5 ms pulse width, and a 5% duty cycle, resulting in an I_SPTA.3_ of 720 mW/cm^2^, and a mechanical index of 0.74 for 10 minutes of stimulation, delivered as 10 30-second trains with a 30 second intertrain interval. Each treatment visit included two sonication blocks. Targeting was guided using either MR guidance and repositioning of the transducer (during the in-scanner visits 1 and 4) or using neuronavigation with BrainSight (during visits 2 and 3), individualized to subject anatomy to ensure accurate focal delivery to each participant’s right amygdala.

### Acoustic Modeling

Acoustic modeling was performed using BabelBrain for participants enrolled at the UCLA site to assess targeting accuracy and provide contextual estimates of target exposure^17^. BabelBrain allows for the simulation of acoustic effects achieved with an ultrasound transducer trajectory (location and orientation of the transducer toward a target region) for an individual brain, considering the type of transducer, the distance from the transducer surface to the scalp, the sonication parameters, and distortion effects caused by the skull (estimated by BabelBrain from the individual T1-weighted data). The 65-mm fixed-focal-length BrainSonix transducer used in this work is supported in BabelBrain. A 3mm gap between the scalp and transducer was included in all simulations to account for the use of acoustic coupling gel pads.

For each participant, we simulated acoustic fields for two transducer trajectories: the position used during the first treatment session, which took place inside the MR environment, and the approximate position used during subsequent sessions, which took place outside the scanner. We normalized each simulated field to its maximum and thresholded it at ≥50% of the maximum simulated pressure. We then averaged the two thresholded maps to create a composite estimate of the suprathreshold beam delivered across treatment sessions. From this composite beam, we calculated five targeting metrics: (1) the percentage of the beam volume that fell within the right amygdala; (2) the percentage of the right amygdala volume covered by the beam; (3) the percentage of the right basolateral amygdala covered by the beam; (4) the percentage of the right centromedial amygdala covered by the beam; and (5) the Euclidean distance between the beam’s peak-pressure point and the center of the right amygdala (MNI coordinates: 22, −8, −24). Individual anatomical images were processed with FreeSurfer^18^, and amygdala nuclei were segmented using FreeSurfer’s amygdala nuclei module^19-21^.

## Results

### Behavioral Findings

For the primary endpoint, HAM-A scores decreased by 10.33 points (SD = 9.51) in the active group and 8.81 points (SD = 9.89) in the sham group between Visits 1 and 4. With this small sample, the change did not significantly differ between groups; mean difference = −1.52 points, 95% CI [−8.65, 5.61], Welch’s t(28.98) = −0.44, p = 0.333, Cohen’s d = −0.16. Using a one-sample t-test (combining active and sham groups), there was an average decrease of 9.55 points (9.58) on the HAM-A between visit 1 and visit 4, with t(30) = -5.55 and a resultant p < 0.001. We used a linear mixed-effects model to examine HAM-A scores across all visits, with fixed effects of visit, treatment group, gender, and site, as well as a visit-by-treatment interaction and a participant-level random intercept. Visit had a significant main effect (p <.001), indicating that HAM-A scores changed over the course of the study. However, the visit-by-group interaction was not significant (p = .934), indicating that HAM-A score trajectories did not differ between the active and sham groups. Figure 1 shows the trajectory of HAM-A scores over time. There were no significant main effects of treatment group (p = .963), gender (p = .456), or site (p = .099).

**Figure 1.**
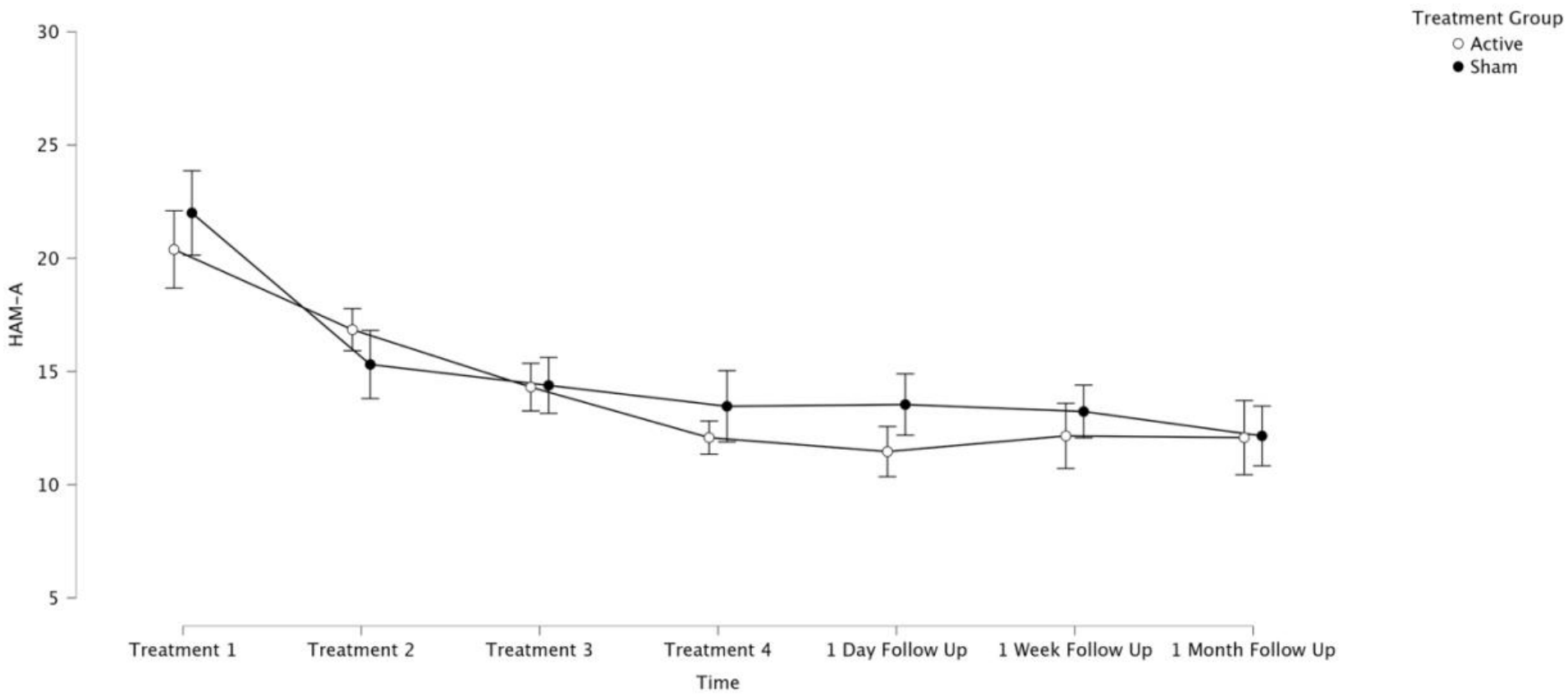
This shows HAM-A scores(+/-SE) between the active and sham arms of the study. Visits 1-4 were treatment visits, while visits 5-7 were follow-up visits.

Further statistical testing is shown below in Table 2.

**Table 1.** Baseline Demographics: Changes in Active vs. Sham Stimulation Groups.

|  | *Active* | *Sham* | *Statistic* | *p-value* |
|---|---|---|---|---|
| *Baseline Demographics* |  |  |  |  |
| Age (n=29) | 42.69 (13.93) | 41.18 (15.35) | t(27)= 0.274 | 0.786 |
| HAM-A at Visit 1 (n=31) | 23 (7.64) | 21.38 (6.88) | t(29)= 0.623 | 0.538 |
| HAM-D at Visit 1 (n=30) | 17.14 (8.53) | 15.18 (6.35) | t(28)= 0.718 | 0.479 |
| GAD-7 at Visit 1 (n=27) | 10.80 (4.86) | 11.42 (5.33) | t(25)= -0.314 | 0.756 |
| PHQ-9 at Visit 1 (n=27) | 11.67 (5.60) | 13.33 (7.58) | t(25)= -0.657 | 0.517 |
| CGI-S at Visit 1 (n=31) | 4 (0.76) | 4.25 (0.45) | t(29)=-1.13 | 0.269 |
| Gender (%female) (n=31) | 86.7% | 50% | χ²(1)= 4.763 | **0.029** |

**Table 2.** Clinical Outcome: Changes in Active vs. Sham Stimulation Groups.

|  | *Active* | *Sham* | *Statistic* | *p-value* | *Effect size* |
|---|---|---|---|---|---|
| *Change from Visit 1 to Visit 4* |  |  |  |  |  |
| HAM-A (n=31) | -10.33 (9.51) | -8.81 (9.89) | t(29)= -0.436 | 0.333 | -0.157 |
| HAM-D (n=30) | -6.29 (9.47) | -4.01 (7.73) | t(28)= -0.708 | 0.242 | -0.259 |
| GAD-7 (n=24) | -5.31 (6.29) | -3.27 (5.53) | t(22)= -0.834 | 0.207 | -0.342 |
| PHQ-9 (n=25) | -2.93 (4.97) | -1.27 (4.41) | t(23)= -0.880 | 0.194 | -0.355 |
| CGI-S (n=29) | -0.73 (1.16) | -1.25 (0.86) | t(29)= 1.42 | 0.916 | 0.509 |
| *Change from Visit 1 to Visit 5* |  |  |  |  |  |
| HAM-A (n=28) | -11.40 (10.39) | -8.46 (9.30) | t(26)= -0.783 | 0.220 | -0.297 |
| HAM-D (n=27) | -8.29 (9.72) | -3.54 (6.37) | t(25)= -1.488 | 0.075 | -0.573 |
| GAD-7 (n=26) | -4.36 (6.13) | -3.50 (4.91) | t(24)= -0.389 | 0.350 | -0.153 |
| PHQ-9 (n=25) | -2.62 (5.88) | -1.92 (5.37) | t(23)= -0.309 | 0.380 | -0.124 |
| CGI-S (n=28) | -0.87 (1.06) | -0.92 (1.04) | t(26)= 0.142 | 0.556 | 0.054 |

Unless otherwise noted, baseline measures used two-tailed tests, while one-tailed tests were used for between-group comparisons based on an a priori directional hypothesis that active stimulation would produce greater improvement than sham. Effect sizes are shown as Cohen’s d for t-test and Cramer’s V for χ^2^ tests. Differences in sample size across analyses reflect the use of available-case data for the primary endpoint and inclusion of all available longitudinal observations in the repeated measures analysis.

Additionally, a responder analysis was performed. Across all responder thresholds tested (25%, 30%, and 35%), there was no statistically significant difference in response rates between the active and sham groups. At the 25% threshold, the sham group had a slightly higher proportion of responders (75% sham vs. 66.7% active), but the chi-square test was non-significant (χ^2^ = 0.261, p = 0.61). At the 30% threshold, the difference was more pronounced (75% sham vs 53.33% active), yet still not statistically significant (χ^2^ = 1.59, p = 0.21). At the 35% threshold, the responder rates were nearly identical (56.25% sham vs. 53.33% active) with a chi-square value of 0.027 (p = 0.87). In all cases, odds ratios were inconclusive with wide confidence intervals crossing 1, and effect sizes were small. These findings suggest no detectable benefit of the active intervention over sham at any responder threshold, though limited sample size may have contributed to insufficient power.

### Blinding Analysis

At the end of visit 1 and visit 4, subjects were queried about which treatment they thought they had received (active vs sham). At visit 1, 33.33% of subjects in the sham group correctly guessed they were in the sham group, while 73.33% of subjects in the active group correctly guessed they were in the active group. The sham group had a Bang’s Blinding Index (BI) of -0.33, indicating a tendency for participants to believe they were receiving the active treatment (suggesting a successful sham). The active group had a BI of 0.47, suggesting partial unblinding. A Fisher’s exact test showed no statistically significant association between the groups (OR = 4.88, p = .066), and the overall correct guess rate across groups (53.3%) did not significantly differ from chance (p = 0.856).

At visit 4, correct group identification was observed in 50% of the sham group participants and 73.33% of those in the active group. The sham group had a Bang’s Blinding Index (BI) of 0, consistent with chance-level identification of treatment allocation. The active group showed a Bl of 0.47, suggesting partial unblinding. A Fisher’s exact test showed no statistically significant association between the groups (OR = 2.56, p = .273), and the overall correct guess rate across groups (61.3%) did not deviate significantly from chance (p = 0.281).

### Modeling Findings

Post hoc subject-specific acoustic modeling in active-treatment participants at the UCLA site (n = 4) evaluated targeting accuracy and associations between target engagement and symptom change.

Simulated acoustic effects achieved in each participant are visualized in Fig. 2. Acoustic modeling demonstrated variable target engagement (see Table 3). On average, 38.21% of the composite suprathreshold beam fell within the right amygdala (SD = 9.52%; range = 30.35–51.68%), while the beam covered only 6.87% of the right amygdala volume (SD = 2.88%; range = 4.19–10.44%). Coverage of the right basolateral amygdala was similar (M = 7.47%, SD = 3.25%; range = 4.64–11.29%), whereas the right centromedial amygdala was not reached by the suprathreshold beam in any participant. The peak-pressure point of the modelled beam lay a mean of 7.54 mm from the center of the right amygdala (SD = 2.18 mm; range = 4.82–9.39 mm).

**Table 3.** Acoustic targeting metrics for active-treatment participants at the UCLA site.

|  | **% beam within R amygdala** | **% R amygdala hit** | **% R basolateral hit** | **% R centromedial hit** | **Peak distance (mm)** |
|---|---|---|---|---|---|
| Group level (n = 4) |  |  |  |  |  |
| Mean | 38.21 | 6.87 | 7.47 | 0.00 | 7.54 |
| SD | 9.52 | 2.88 | 3.25 | 0.00 | 2.18 |
| Median | 35.40 | 6.43 | 6.98 | 0.00 | 7.98 |
| Range | 30.35–51.68 | 4.19–10.44 | 4.64–11.29 | 0.00–0.00 | 4.82–9.39 |
| Individual participants |  |  |  |  |  |
| 05 | 37.94 | 7.95 | 9.04 | 0.00 | 9.22 |
| 06 | 30.35 | 4.19 | 4.64 | 0.00 | 9.39 |
| 10 | 32.87 | 4.92 | 4.92 | 0.00 | 6.74 |
| 12 | 51.68 | 10.44 | 11.29 | 0.00 | 4.82 |

*Note. Values are the mean of the in-scanner (first treatment) and out-of-scanner (subsequent treatments) simulations. Percentages refer to the suprathreshold beam (≥50% of the maximum simulated pressure). Peak distance is the Euclidean distance from the beam's peak-pressure point to the centre of the right amygdala. SD = sample standard deviation.*

**Figure 2:**
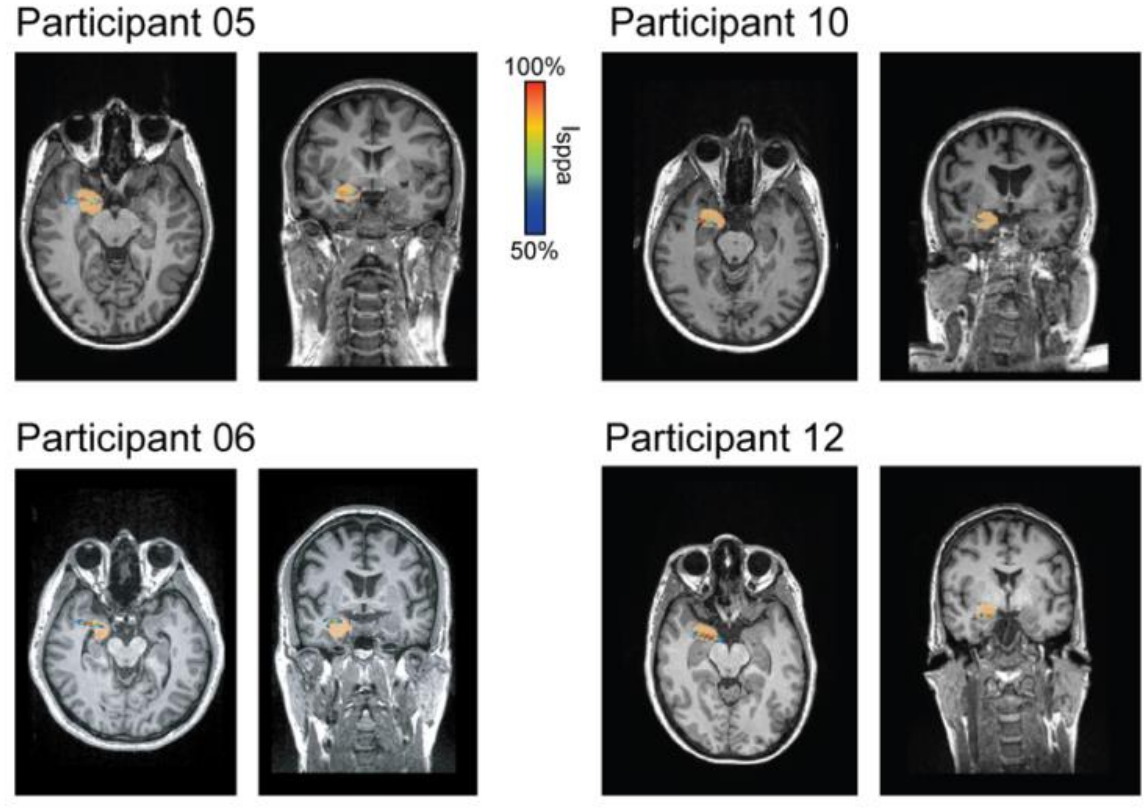
Simulating acoustic effects of targeting right amygdala in active treatment participants at the UCLA site. The right amygdala is colored orange in all participants. The focus of the simulated ultrasound beam is colored by the estimated acoustic pressure, ranging from 50% (dark blue) to 100% (dark red) of the individual’s maximum pressure achieved.

Associations between target engagement and symptom change were examined using two targeting metrics, the percentage of the right amygdala covered by the suprathreshold beam and the distance from the beam’s peak-pressure point to the center of the right amygdala, each averaged across the in-scanner and out-of-scanner simulations (Fig. 3a & b). Across both intervals, greater amygdala engagement tended to be associated with less improvement on the HAM-D, GAD-7 and PHQ-9. The strongest associations were between amygdala coverage and PHQ-9 change, from Visit 1 to Visit 4 (r = 0.99, p = .01) and from Visit 1 to Visit 5 (r = 0.99, p = .01), as well as between peak distance and GAD-7 change from Visit 1 to Visit 4 (r = −0.97, p = .03), where a shorter distance to the amygdala center was associated with less improvement. None of these associations survived Bonferroni correction for multiple comparisons (20 tests; corrected α = .0025; smallest adjusted p = 0.12). Given the small sample, these analyses are descriptive.

**Figure 3.**
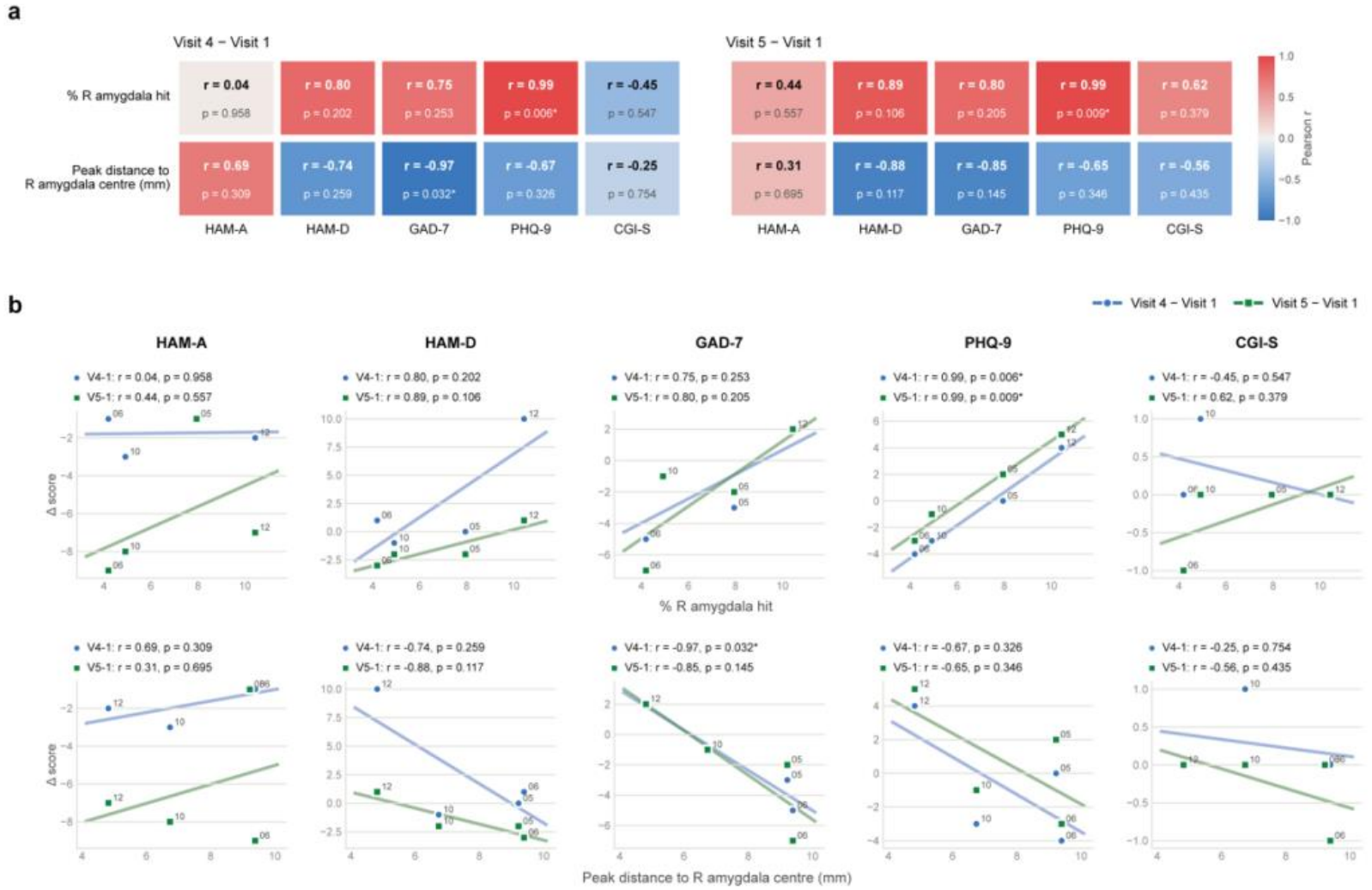
Associations between acoustic target engagement and symptom change amongst active-treatment participants at UCLA site (n = 4). **(a)** Pearson correlations (r) and p-values between each targeting metric and change in HAM-A, HAM-D, GAD-7, PHQ-9 and CGI-S scores from Visit 1 to Visit 4 (left) and Visit 1 to Visit 5 (right). **(b)** Scatter plots of each targeting metric against symptom change from Visit 1 to Visit 4 (blue circles) and from Visit 1 to Visit 5 (green squares), with least-squares regression lines. Points are labelled by participant number. *p < .05, uncorrected

**Figure 4.**
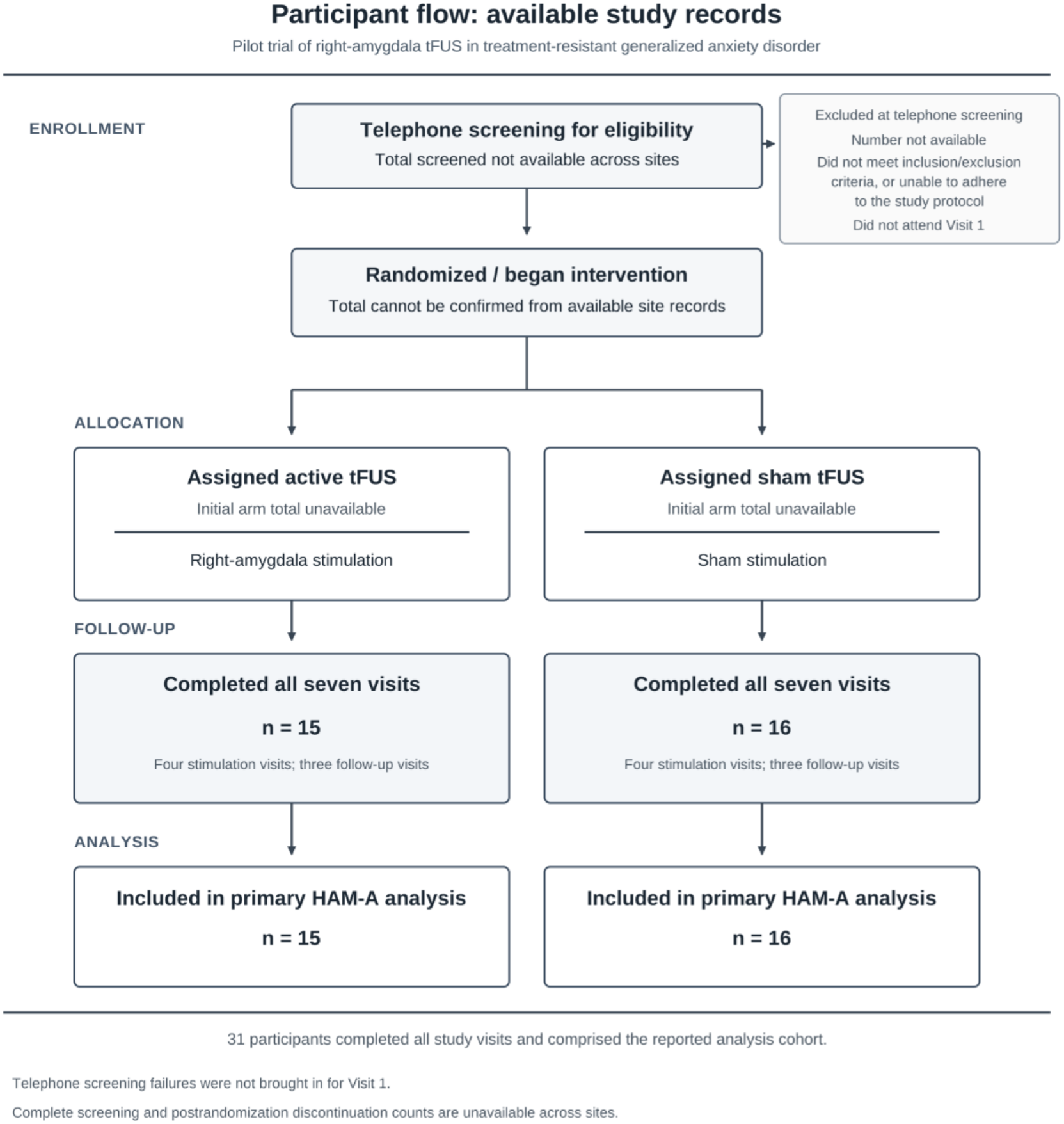
CONSORT flow diagram.

## Discussion

In this double-blind, sham-controlled pilot trial, active tFUS targeting the right amygdala did not result in significantly greater improvement in anxiety symptoms compared to sham stimulation. Although both groups showed substantial reductions in HAM-A scores over time, we observed no significant between-group differences on the primary endpoint or across secondary clinical measures, though the active group had larger decreases in HAM-A between visit 1 and visit 5 (p=0.22, Cohen’s d= -0.297).

Based on this effect size, 282 patients would be needed to achieve 80% power to detect such an anxiolytic effect. Notably, this is on average less than the sample size needed for SSRI medications, which can range from 216 (for sertraline) to 620 (for paroxetine) ^22^.

The change in Hamilton Depression Rating Scale (HAM-D) scores from visit 1 to visit 5 showed a non-significant difference (p=0.078) between active and sham, with a moderate effect size (Cohen’s d = -0.57). Although the effect was not statistically significant, its size indicates a possible signal that merits further study in larger trials. There is a potential for moderate antidepressant effects of amygdala tFUS in anxiety groups, aligning with results from other amygdalar tFUS studies in depression populations ^24^.

Nevertheless, estimating from this effect size, about 76 patients would be needed to achieve 80% power to detect such an antidepressant effect. Since this study involved only 31 patients, it was severely underpowered to detect small-to-moderate treatment effects in either depression or anxiety ratings.

At baseline, the groups did not differ significantly except for gender: the active group was 86.7% female, and the sham group was 50% female. Although the overall study population is 67.74% female– consistent with the lifetime prevalence of GAD being about twice as high in women as in men–the gender disparity in randomization may have introduced unaccounted variability in treatment response and should be considered a limitation ^23^.

Blinding efficacy was assessed by asking participants to guess their group assignment immediately following treatment at both visits 1 and 4. Analysis of visit 1 guesses showed no significant difference in guess accuracy between groups, and overall guesses did not deviate from chance, suggesting adequate initial blinding. Although the active group demonstrated a BI of 0.47, indicative of moderate unblinding with a tendency toward correct identification, this finding did not reach statistical significance and may reflect variability related to the small sample size rather than true unblinding.

At visit 4, blinding efficacy remained largely unchanged in the active group. In the sham group, the BI shifted to 0.00, indicating perfect blinding at the group level. However, this change also reflects a redistribution of guesses, with a greater proportion of sham participants endorsing the belief that they were receiving active treatment. This pattern may reflect expectation-related effects or perceived engagement with the intervention over time. Alternatively, it may reflect random variation given the limited sample size. These findings should therefore be interpreted cautiously, and no definitive conclusions should be drawn about changes in blinding over time.

Neuroimaging analyses did not demonstrate a significant effect of active tFUS relative to sham. Neither the perfusion-based analyses nor the simultaneous BOLD analyses identified significant between-group differences, and the resting-state functional connectivity analyses similarly failed to show a significant active-versus-sham effect, including when the analysis was restricted to the right prefrontal cortex. The within-group changes in BOLD signal in the default mode network and functional connectivity warrant cautious interpretation, as they were not supported by significant differences between active and sham treatment. Accordingly, these findings cannot be attributed specifically to ultrasound stimulation and should not be interpreted as evidence of target-specific neuromodulation.

Taken together, the imaging results provide no clear evidence of differential neural engagement by active tFUS under the stimulation parameters used in this study^25^.

Furthermore, there was no reappraisal or cognitive component prior to stimulation. Published literature in psychiatric neuromodulation typically has a component where subjects are exposed to their psychological stressors before receiving neuromodulation to downregulate the created distress^12,26^. This is also now the case with psychedelic-assisted psychotherapy (such as ketamine)^27^. The lack of a cognitive component in the stimulation done in this study may be partly responsible for the lack of separation between the active and sham arms.

Another likely limitation in this study was the intensity of stimulation used. This study administered tFUS with an I_SPTA.3_ = 720 mW/cm^2^, in line with the FDA guidelines for diagnostic ultrasound, which has dictated the intensity for most neuromodulation studies using tFUS. This may have led to insufficient energy deposition at the amygdala, thereby limiting the neuromodulatory effect. This is a commonly proposed issue of the tFUS intensity guideline, especially given the role of hair and skull in sonication aberration and attenuation^28,29^.

Several studies over the last few years have demonstrated the safety of intensities beyond the FDA guideline ^30-33^, paving the way for clinical neuromodulation trials to investigate the possible increased clinical effect associated with higher energy deposition. We believe this may be especially useful in GAD and other anxiety disorders, in which substantial placebo responses can obscure treatment effects in early-stage neuromodulation trials. Increasing tFUS intensity may help overcome this by producing more robust neuromodulatory engagement of target circuits, thereby improving the likelihood of detecting clinically meaningful differences in behavioral scales and neuroimaging changes between active and sham conditions.

The convergence of the behavioral, neuroimaging, and acoustic modeling findings suggests that the failure to detect a statistically significant treatment effect may have reflected the small sample size as well as insufficient neuromodulatory engagement rather than failure to target the intended anatomy.

Active stimulation did not separate from sham on the primary clinical endpoint or on responder analyses, and no significant stimulation-related effects were detected on simultaneous BOLD or perfusion imaging at the prespecified thresholds. In contrast, subject-specific acoustic modeling in the UCLA cohort demonstrated that the simulated ultrasound field reached and overlapped the right amygdala, supporting the plausibility of anatomical targeting. Taken together, these findings raise two related possibilities: the delivered acoustic dose may have been insufficient to produce a reliable physiological effect at the target, or stimulation of the amygdala in the absence of concurrent emotional or cognitive engagement may not have been an optimal paradigm for producing measurable circuit or behavioral change. The present results demonstrate the feasibility of amygdalar tFUS and support future studies incorporating higher-energy exposure, refined dosing, and state-dependent or symptom-provocation paradigms designed to engage the targeted circuit during sonication.

## Conclusion

This pilot randomized, double-blind, sham-controlled trial demonstrates the feasibility of targeting the right amygdala with tFUS in patients with treatment-resistant GAD, but did not demonstrate a statistically significant benefit over sham in this small sample at the stimulation parameters tested. Active and sham groups showed reductions in anxiety symptoms over time, but there was no statistically significant between-group separation on the primary clinical endpoint or secondary behavioral measures. Subject-specific acoustic modeling demonstrated that the simulated ultrasound field reached and overlapped the intended right amygdala target, supporting the plausibility of anatomical targeting despite the absence of measurable physiological or clinical effects.

Future trials should therefore evaluate higher acoustic exposures, various pulse repetition and carrier frequencies, length of stimulus trains, frequency of stimulation, refined dose-response relationships, and state-dependent or symptom-provocation paradigms while incorporating larger samples adequately powered to detect clinically meaningful active-versus-sham differences. These results establish that the stimulation parameters and paradigm tested here were feasible but did not produce detectable differential clinical effects.

## Data Availability

All data produced in the present study are available upon reasonable request to the authors

## Acknowledgments

Tiny Blue Dot, MSG support by the Joseph Loveless Foundation. Dr. George was supported, in part, by the National Institute of General Medical Sciences of the National Institutes of Health under Grant Number P30GM154630. Support from DOD CDMRP HT9425-24-1-1081 (to MMM) & NIH R21EB034428 (to MMM). NIA R01AG073480 (to SYB). F30MH136802 (to NMS). T32GM152342 (to NMS)

## Notes

### Clinical Trial

NCT04557891

### Author Declarations

The study was registered on ClinicalTrials.gov (NCT04557891) before the first patient was enrolled. Patients with treatment-resistant GAD were enrolled across 3 sites (University of California, Los Angeles (UCLA), Massachusetts General Hospital (MGH), and Medical University of South Carolina (MUSC)). UCLA served as the data coordinating center, and the UCLA Medical IRB reviewed the study (IRB: 20-1750); sites also maintained local IRB approvals.

